# The assessment of Computer Vision Algorithms for the Diagnosis of Prostatic Adenocarcinoma in Surgical Specimens

**DOI:** 10.1101/2020.07.14.20152116

**Authors:** Syed Usama Khalid Bukhari, Ubeer Mehtab, Syed Shahzad Hussain, Asmara Syed, Syed Umar Armaghan, Syed Sajid Hussain Shah

**Affiliations:** Assistant Professor, Department of Computer Science, The University of Lahore, Islamabad - Pakistan; House officer, Bahawal Victoria Hospital, Quaid e Azam Medical College, Bahawalpur - Pakistan; Lecturer, Electrical Engineering Department. National University of Technology (NUTECH), Islamabad, Pakistan; Lecturer, Department of Pathology, Faculty of Medicine, Northern Border University, Arar-Kingdom of Saudi Arabia; Student, Biomedical Engineering, Riphah International University. Islamabad - Pakistan; Professor, Department of Pathology, Faculty of Medicine, Northern Border University, Arar-Kingdom of Saudi Arabia

**Keywords:** Computer Vision, Deep Learning, Prostatic Carcinoma, ResNet

## Abstract

**Introduction:** Prostatic malignancy is a major cause of morbidity and fatality among men around the globe. More than a million new cases of prostatic cancer are diagnoses annually. The incidence of prostatic malignancy is rising and it is expected that more than two million new cases of prostatic carcinoma will be diagnosed in 2040. The application of machine learning to assist the histopathologists could be a very valuable adjunct tool for the histological diagnosis of prostatic malignant tumors.

**Aim & Objectives:** To evaluate the effectiveness of artificial intelligence for the histopathological diagnosis of prostatic carcinoma by analyzing the digitized pathology slides.

**Materials & Methods:** Eight hundred and two (802) images in total, were obtained from the anonymised slides stained with hematoxylin and eosin which included anonymised 337 images of prostatic adenocarcinoma and 465 anonymised images of nodular hyperplasia of prostate. Eighty percent (80%) of the total digital images were used for training and 20% for testing. Three ResNet architectures ResNet-18, ResNet-34, and ResNet-50 were employed for the analysis of these images.

**Results:** The evaluation of digital images by ResNet-18, ResNet-34, and ResNet-50 revealed the diagnostic accuracy of 97.1%, 98 % and 99.5 % respectively.

**Discussion:** The application of artificial intelligence is being considered as a very useful tool which may improve the patient care by improving the diagnostic accuracy and reducing the cost. In radiology, the application of deep learning to interpret radiological images has revealed excellent results. In the present study, the analysis of pathology images by convolutional neural network architecture revealed the diagnostic accuracy of 97.1%, 98 % and 99.5 % with by ResNet-18, ResNet-34, and ResNet-50 respectively. The findings of the present study are in accordance with the other published series, which were carried out to determine the accuracy of machine learning for the diagnosis of cancers of lung, breast and prostate. The application of deep learning for the histological diagnosis of malignant tumors could be quite helpful in improving the patient care.

**Conclusion:** The findings of the present study suggest that intelligent vision system possibly a worthwhile tool for the histopathological evaluation of prostatic tissue to differentiate between the benign and malignant disorders.

## Introduction

Prostatic carcinoma is the second most prevalent type of malignant tumor diagnosed in male all over the world (1). In 2018, more than one million (1,276106) patients have been diagnosed as a case of prostatic cancer around the globe and this malignant tumor caused 358989 deaths during that time period (1).

Variation in the prevalence of cancers exists among the different geographic regions (2). Some tumors are more prevalent in one region and less common in other. The prostatic cancer is the most frequent malignant tumor of male population in one hundred and five countries which mainly include the countries from the continents of Europe, Africa, North & South America (3). The geographic variation in the prevalence of prostatic cancer has been attributed to genetic and environmental factors. Higher consumption of meat has been linked to higher risk of development of prostatic carcinoma (4-5).

The incidence of prostatic malignancy rises with increase in the age. The prostatic malignancy is more common in 7^th^ decade with mean age of sixty-six years (3). An increased incidence of prostatic adenocarcinoma has been observed in the younger population. This rising trend in the incidence of cancer of prostate in the younger age group has been attributed to improved diagnostic techniques and environmental factors including the rising incidence of obesity among the adolescent and younger age groups (6).

The prostatic carcinoma causes urinary tract symptoms due to enlargement of prostrate. The spread of prostatic adenocarcinoma to vertebral column causes backache. The common symptoms with which these patients presents usually includes urinary obstruction, urgency, hesitancy, discomfort, frequency and hematuria (7-9). The suspected cases of prostatic malignancy are tested for prostate specific antigen. There is an increased suspicion of prostatic carcinoma in case of elevated blood PSA level. But the definitive diagnosis of prostatic malignant tumor is confirmed by the histopathological examination of tissue specimen from the prostate. The rising number of prostatic biopsies for the histopathological examination are going to increase the stress on the histopathologists in future as more than two million cases of prostatic cancer are expected by year 2040 (3).

The application of artificial intelligence and development of Computer aided diagnostic programs may be instrumental in enhancing the laboratory efficiency by reducing the turnaround time, cost and chance of human errors. Artificial intelligence is yielding very good results in the interpretation of digital images by analyzing them with Deep learning architecture.

The main focus of this study is to assess the usefulness of machine learning for the histological diagnosis of prostatic adenocarcinoma.

## Materials & Methods

The research project has been approved from the ethical committee of University of Lahore - Islamabad Campus. Eight hundred and two (802) digital anonymized images were collected, which included 337 images of prostatic adenocarcinoma and 465 images of prostatic hyperplasia. We have used three ResNet convolutional neural network architectures, ResNet-18, ResNet-34 and ResNet-50. Because of dataset limitation, the transfer learning approach has been employed to train the models using the ImageNet data set (10).

A total of 337 images of prostatic adenocarcinoma and 465 images of nodular hyperplasia of prostate are used in the present research project. These images have been reviewed and labeled by two histopathologists.

A random approach of FastAI API is used to load the data into train and test sets, to avoid the chances of images of the same class been selected for a mini-batch input, that can harm the performance of model validation.

Two sets (training set and test set) are made from the whole dataset. The training set is comprised 80% of total dataset, while the Testing set is 20% of total dataset.

For image, we used data augmentation for regulation.

## Results

A total of eight hundred and two (802) images were collected. Three hundred and thirty-seven digital images (337) of adenocarcinoma of prostate and four hundred and sixty-five (465) images of prostatic hyperplasia. Six hundred and forty (640) images randomly chosen from both carcinoma and prostatic hyperplasia images that is 80% of the total data set, and employed for training data set. Where one hundred and sixty-two (162) images randomly chosen from both carcinoma and hyperplasia images that is 20% of the total data set employed in the test data set.

Three ResNet architecture models, ResNet-18, ResNet-34 and ResNet-50 are used to classify the input image. Accuracy of 97.1%, 98 % and 99.5 % respectively was obtained on test data set.

**Figure 1:**
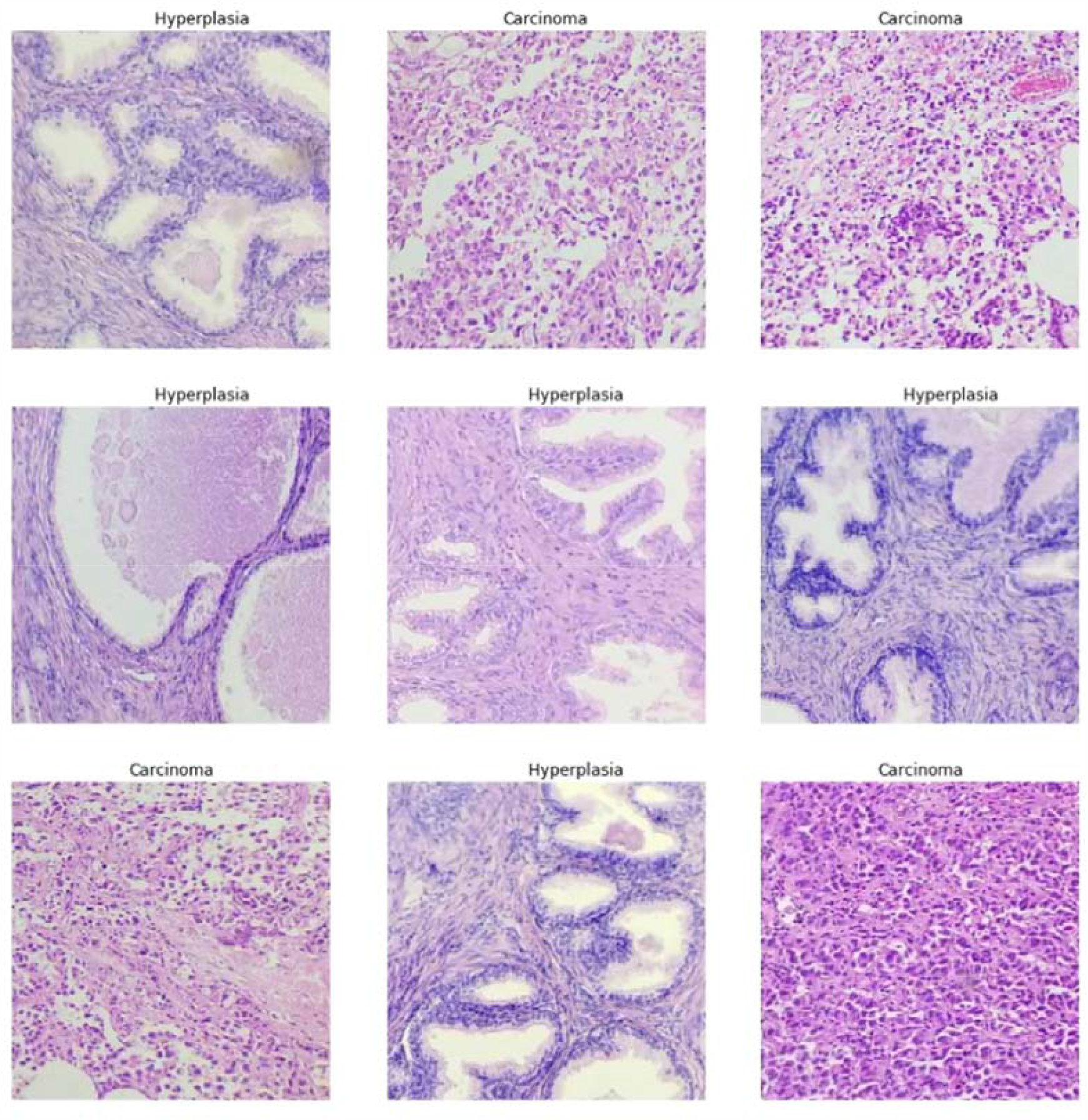
Example of Data and labels loaded using FastAI (randomly)

**Table 1.**
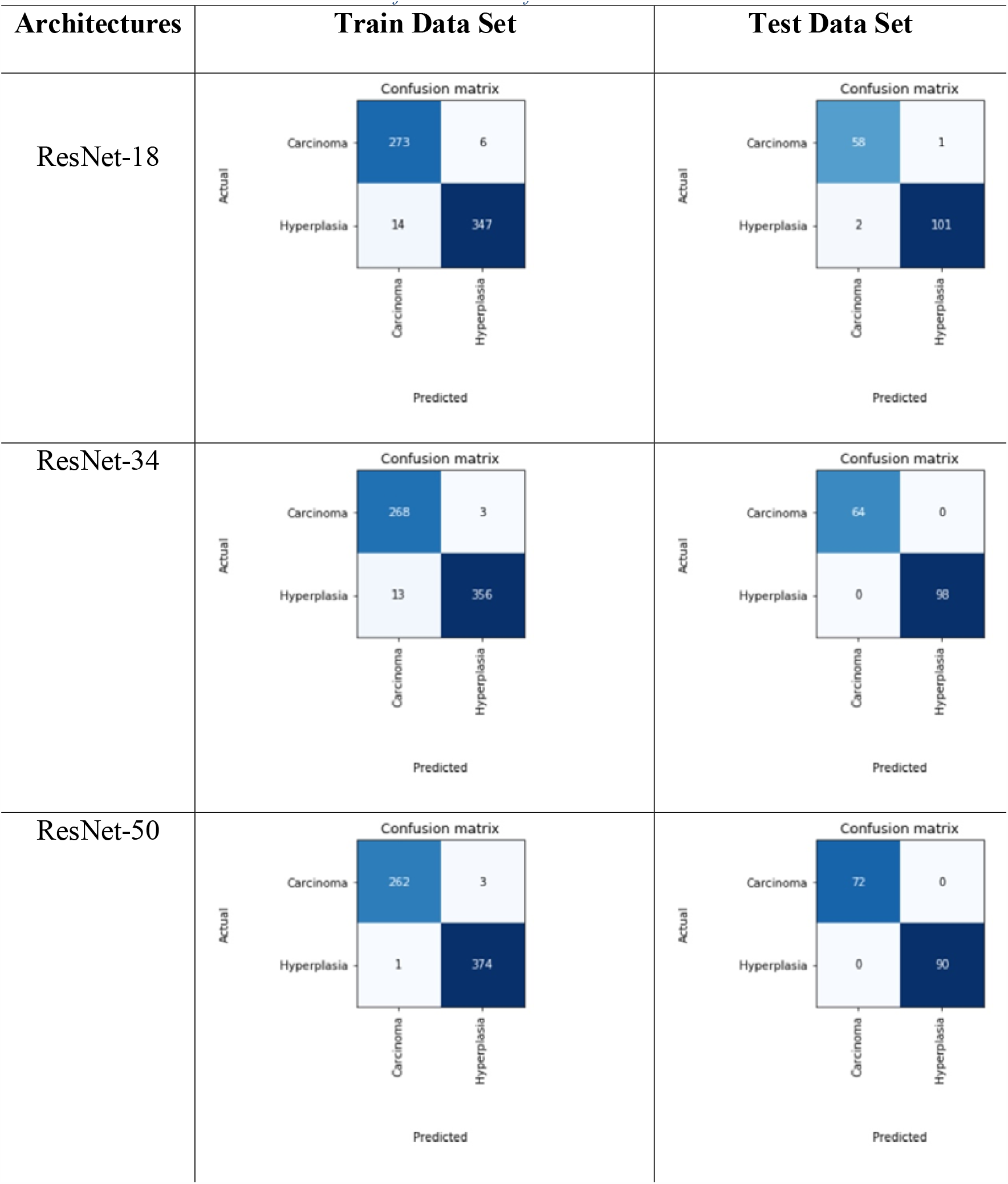
Confusion matrix of train and test datasets.

**Table 2.** Detail results Test data set.

| ARCHITECTURES | PRECISION | RECALL | F-1 SCORE | ACCURACY |
| --- | --- | --- | --- | --- |
| **RESNET-18** | 0.966 | 0.983 | 0.974 | 97.1% |
| **RESNET-34** | 1.0 | 1.0 | 1.0 | 98.0 % |
| **RESNET-50** | 1.0 | 1.0 | 1.0 | 99.5 % |

## Discussion

Adenocarcinoma of prostrate is a quite prevalent tumor in the majority of the countries of the world. More than one million cases of prostatic cancer are diagnosed every year around the globe. Approximately two million persons may become victim of prostatic malignancy every year by year 2040 (3). The rising number of prostatic biopsies for the purpose of histopathological diagnosis is going to mount the burden on the diagnostic laboratories and will increase the stress on the histopathologists. In order to overcome this situation, the application of smart technology such as artificial intelligence may be very helpful.

The rapid advancement in field of artificial intelligence in the recent past has drawn the attention of health care providers to apply this technology for the solution of complex medical problems.

The artificial intelligence is characterized by development of machines which have the capability to do cognitive assignments for the achievement of desirable output based on given input. The application of artificial intelligence has been expected to revolutionize the patient care by improving the diagnostic accuracy and reducing the cost with better choices for the management of diseases (11). An important sub group of artificial intelligence is Deep learning. The application of deep learning revealed very encouraging results in the field of radiology for image interpretation. A study conducted by Pasa F et al showed very encouraging results of Convolutional neural network (CNN) for the image analysis of the X-rays. They used convolutional neural network to diagnose tuberculosis (12).

The histopathological diagnosis of malignant tumors is based upon certain specific microscopic features such as pleomorphism, hyperchromasia, high nucleus to cytoplasmic ratio, atypical mitosis, necrosis, multinucleated giant cells and pattern of arrangement of neoplastic cells. The assessment of digital pathology image with machine learning may provide valuable assistance in the interpretation and differentiation of neoplastic and non-neoplastic lesions.

The convolutional neural network has been applied in this research project to diagnose prostatic adenocarcinoma by analyzing the digital pathology images which revealed excellent results. With the application of ResNet-50 architecture, a diagnostic accuracy of 99.5 % has been achieved while ResNet-34 and ResNet-18 yielded diagnostic accuracy of 98 % and 97.1%, respectively. The findings of the present study are in accordance with the other published series which were carried out to assess the accuracy of deep learning for the diagnosis of cancers of lung, breast and prostate (13-16).

The histopathological assessment is quite subjective matter which may be a possible reason for the inter-observer discrepancies. The advancement in the computational image technology may be able to make the histopathology more objective and quantitative (17).

The application of computer based diagnostic system in histopathology will be a great help to the histopathologists in the current scenario of rising trend in the incidence of neoplastic lesions leading to increase in the number biopsies.

## Conclusion

The findings of the present research project raises the potential application of computer vision-based systems as a valuable tool for the histopathological evaluation of prostatic tissue in differentiating the benign and malignant disorders. The use of intelligent vision system in the field of histopathology for the diagnosis of malignant disorders may also, lessen the risk of human errors and it will decrease the workload on histopathologists along with provision of a second opinion to them.

## Data Availability

NA

## Acknowledgement

The authors are grateful to Farhan Mazher for his valuable assistance.

